# Exploring measures to increase detection of malaria cases through reactive case detection in a Southern Province of Zambia-like Setup: A modelling study

**DOI:** 10.1101/2024.07.18.24310660

**Authors:** Chilochibi Chiziba, Sheetal Silal

**Affiliations:** Modelling and Simulation Hub, Africa, Department of Statistical Sciences, University of Cape Town, Cape Town, South Africa; Centre for Global Health, Nuffield Department of Medicine, Oxford University, Oxford, United Kingdom

## Abstract

**Introduction:** In Zambia, malaria prevalence varies spatially, posing challenges for intervention strategies. Asymptomatic and clinical carriers not accessing healthcare further complicate efforts, necessitating reactive case detection (RCD) to target undetected infections. However, operational hurdles, such as resource shortages and logistical complexities—including shortages of community health workers (CHWs), difficulties reaching residents, and limitations in malaria rapid diagnostic tests (RDTs)—hinder RCD’s effectiveness. Identifying effective improvement measures given circumstances that may lead to deficient intervention outcomes may improve the situation.

**Methods:** A mathematical model of malaria transmission conforming to Zambia’s low transmission areas defined as areas with an incidence of fewer than 200 malaria cases per 1,000 population per year was developed to simulate RCD using parameters and data from published articles. We explored the impact of literature-identified challenges on RCD performance in malaria detection and potential strategies to enhance detection rates. The examined factors and improvement measures included increasing CHWs, adjusting reaction time, RDT sensitivity, and implementing focal mass drug administration (fMDA).

**Results:** Simulation findings suggest that a shortage of CHWs and limited availability of RDTs have the highest negative impact on RCD compared to other challenges. In scenarios where CHWs or RDT availability for RCD were reduced by 50%, annual malaria cases were predicted to increase by approximately 17%. Only the incorporation of fMDA as an improvement measure succeeded in countering the situation. Increasing CHWs to offset RCD inefficiencies caused by limited RDT sensitivity or difficulties in finding individuals resulted in fewer cases than improving reaction time or increasing the screening radius.

**Conclusions:** Participation of CHWs is voluntary and primarily motivated by informal incentives, often provided by donors. Finding sustainable means to ensure the sufficient availability of CHWs may guarantee continued RCD contributions towards maintaining stable malaria prevalence and elimination. More research is required to explore the application of RCD in archetypical transmission areas suitable for RCD as improvement measures to the identified challenges hindering RCD. Furthermore, archetype-based targeting of interventions, including RCD, may also be explored to inform the optimisation of intervention resource allocation to overcome the widening gap in malaria funding.

## Introduction

Malaria has proved difficult to eradicate despite being treatable, likely due to its complex transmission dynamics [1]. Although some countries have successfully eliminated malaria, countries in sub-Saharan Africa still carry the highest burden, accounting for over 90% of the 247 million cases reported globally [2]. Furthermore, the malaria burden varies significantly across sub-Saharan Africa and within countries, including Zambia [2,3]. Complicating the implementation of intervention strategies are the complexities of transmission and heterogeneity as well as the presence of individuals who fail to seek treatment despite exhibiting symptoms and those who remain asymptomatic yet capable of transmitting the disease [4,5].

Although Zambia is not among the top contributors to global malaria cases, as of 2021, 57% of its population lived in regions classified as having low to very low malaria risk (less than 200 cases per 1,000 people per year) [3]. However, malaria risk and malaria parasite prevalence exhibit spatial heterogeneity across smaller spatial boundaries, such as health facility catchment areas (HFCAs)[3,6]. From 2017 to 2021, the distribution of cases per 1000 people per year at HFCA level remained relatively stable, except for a notable increase in high-risk areas (> 500 cases per 1000 per year) from 24% in 2019 to 47% in 2020 [3]. As Zambia works to increase the number of low malaria-risk HFCAs and “to increase malaria-free HFCAs from 10 to at least 250 by 2026” through various interventions, the potential for a resurgence of cases in these regions remains high [5]. This is because the proportion of asymptomatic individuals with low parasite densities among the infected population rises, as malaria transmission rates decrease [5]. Despite being less infectious than symptomatic cases, these individuals form an asymptomatic reservoir capable of transmitting parasites in areas where vectors are present [5]. To combat the resurgence of cases, interventions including reactive case detection (RCD) are strategically implemented to target asymptomatic infections and symptomatic individuals not seeking treatment offering treatment to halt transmission without the need for universal testing or treatment [3,5,7].

The existing Zambia-specific research indicates that RCD is effective in several ways but also impeded by various challenges. A mathematical modelling study conducted by Gerardin et al. (2017) used household locations, demographics, and malaria prevalence data to train an agent-based model to assess the effectiveness of RCD based on different transmission profiles, which included, “low-transmission, high household density; high-transmission, low household density; and high-transmission, high household density [8].” The simulation findings estimated that RCD is only effective in areas that have newly become low transmission areas [8]. Also, Chitnis et al. (2019), in a theoretical modelling paper that used Zambian data found that it is more important to increase the number of index cases followed than to increase the number of neighbours tested per index case, if RCD is to be effective [9]. Similarly, Reiker et al.’s (2019) mathematical modelling study suggests that RCD is ideal in areas where transmission is initially low, and that increasing radius yields relatively better case detection [10]. Furthermore, Larsen et al. (2017) and Bhondoekhan et al. (2020) suggest that prioritizing locations with high environmental susceptibility to malaria transmission during RCD operations is crucial in detecting cases in low transmission areas [4,11]. Additionally, all studies on RCD in Zambia agree that RCD’s efficacy can be improved and that, on its own, it may not lead to malaria elimination in low transmission areas. However, if complemented with other interventions such as reactive focal drug administration (fMDA), it may realistically lead to elimination [4,9–14]. Most importantly, Chitnis et al. (2019) and Reiker et al. (2019) conclude that prevalence reduction due to RCD is mainly determined by the proportion of all infections identified within a specific timeframe [9,10].

While the studies on RCD in Zambia provide valuable insights, recent advancements in malaria interventions will affect RCD outcomes differently, such as advancements in malaria rapid diagnostic test (RDT) sensitivity. Some of these studies also compare the circumstances/settings in which RCD is most efficient. However, their applicability for informing operational decision-making may be limited, considering the operational challenges that impede RCD implementation in resource-constrained settings. These challenges frequently result in relatively fewer detections by RCD, further reducing its effectiveness [5]. An evaluation conducted by Searle et al. (2016) in the low-transmission regions of the Southern Province of Zambia highlighted several operational hurdles hindering the implementation of RCD. These hurdles included inadequate supplies of RDTs, a shortage of community health workers (CHWs), logistical complexities, difficulties in reaching residents in designated households, and the limited sensitivity of RDTs. [5]. These challenges directly impact the proportion of all undetected infections identified through RCD [5]. Given the competing priorities faced by implementers of malaria interventions, including RCD, knowing which improvement measures to undertake given circumstances that may lead to deficient outcomes of an intervention may be improve the situation.

The purpose of this study was to use mathematical modelling to investigate the impact of various literature-informed challenges affecting RCD to reduce malaria cases. This investigation also considered their respective potential improvement measures. These measures include increasing the CHW workforce, improving reaction time, changing diagnostics tests, or pivoting fMDA. These are targeted for the most common situations that lead to inefficient implementation of RCD in low transmission areas in Zambia to relatively better inform operational decisions.

Our study is based on hypothesized RCD conditions that most low transmission HFCAs experience in Zambia, using data from the Southern Province. Hence, the study bears the ’Zambia-like’ conceptualization, meaning it can generally represent any area with similar conditions [5].

## Methods

### Study site

In this study, we simulate a single hypothetical low-transmission HFCA. Specifically, the study used parameters from randomized control trials and cross-sectional studies conducted in low transmission areas of the Southern Province in the years 2014 to 2018 [4,5,10,11,13–16]. In Zambia, RCD is currently implemented in low transmission areas as stratified by HFCA [3]. In such areas, a positive malaria case at the health facility or post triggers an RCD investigation, which is carried out by CHWs assigned to the health post near the index case. One HFCA serves approximately 10,000 people, while a health post, a subset of the HFCA, serves between 500 to 1,000 people [4,5,15]. The

### Malaria reactive case detection model

To mimic the operation of RCD at the HFCA level, a deterministic non-linear ordinary differential equation (ODE) model was developed to simulate malaria transmission and RCD implementation visualized in Figure 1. The figure provides an overview of the RCD malaria transmission model, generalises susceptible exposed infected recovered (SEIR) model format with added treatment compartments. The model includes the human population only with vector dynamics folded into the human force of infection. In this model (Figure 1), individuals progress through distinct compartments representing various stages of infection and treatment. Initially, individuals are categorized as susceptible (S), signifying their vulnerability to malari acquisition. Following exposure to the malaria parasite, individuals transition to the exposed (E) compartment, indicative of infection without immediate infectiousness. Subsequently, individuals may progress to either the asymptomatic (A) or symptomatic (C) compartments, contingent on the manifestation of malaria symptoms. Those symptomatic individuals may undergo therapeutic intervention at a health facility, leading them to the treatment (X) at the health facility or treatment through RCD (V) compartments. Furthermore, some of the asymptomatic (A) individuals may also receive treatment through the screening compartment of RCD compartment V or recover naturally. Ultimately, individuals in the treatment compartment V and X and asymptomatic individuals not treated through RCD advance to the recovered (R) compartment, reflecting either clearance of the infection or the establishment of partial immunity. Tables 1 summarize the parameter definitions that govern the transitions between compartments depicted in Figure 1.

**Figure 1:**
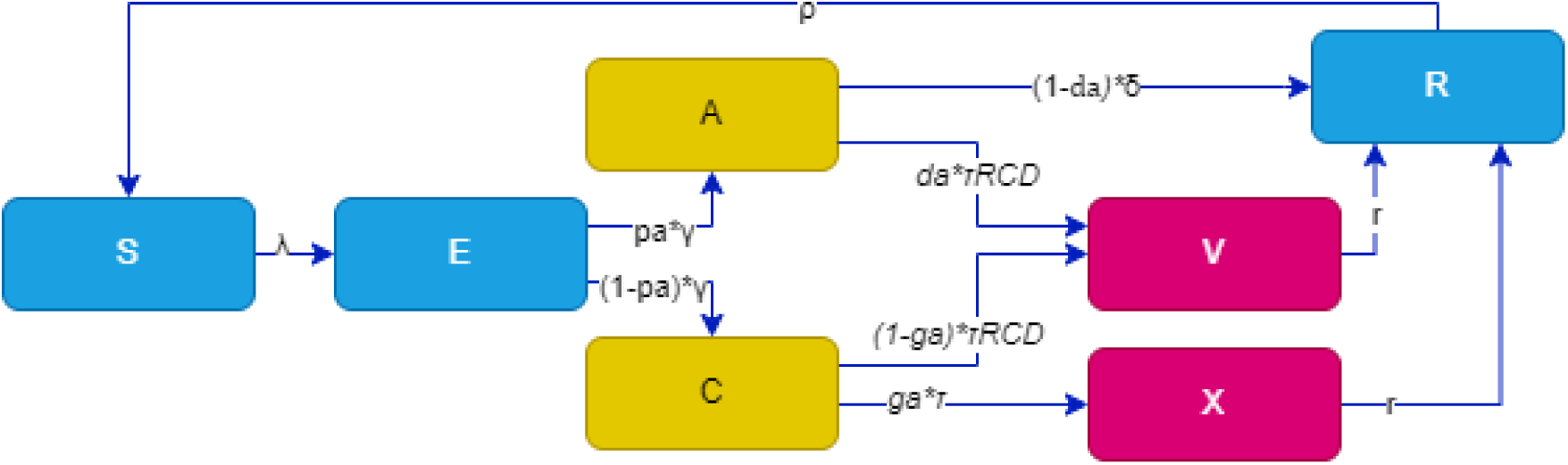
RCD model flow diagram with compartments S (Susceptible), E (Exposed), A (asymptomatic), C (Symptomatic/clinical), V (treatment through RCD), X (treatment at health facility), and R (recovered). The description of parameters governing movements through the compartments are described in Table 1.

**Table 1.** Model parameters, values, descriptions, and sources.

| Symbol | Definition | Value | Range | Source |
| --- | --- | --- | --- | --- |
| **a** | Human feeding rate per mosquito (per day) | 0.33 | (0.10, 1) | [18] |
| **b** | Transmission efficiency from mosquito to human (per day) | 0.022 | (0.010,0.27) | [18] |
| **c** | Transmission efficiency human to mosquito (per day) | 0.24 | (0.072, 0.64) | [18] |
| **pa** | Proportion of asymptomatic infections | 0.395 | (0.2,0.49) | [15] |
| **da** | Proportion of asymptomatic infections that get screened and treated through RCD | 0.22 | (0.15,0.3) | [5] |
| **ga** | Proportion of clinical infections that get treatment at health facility | 0.97 | (0.84,0.99) | [19] |
| $\gamma$ | Rate of onset of infectiousness in humans (Incubation rate; per day) | 0.071 | (0.06,0.08) | [20,21] |
| **$\tau_{RCD}$** | Rate at which infectious population is screened and treated via RCD (per day) | Estimated during analysis |  |  |
| $\lambda$ | Force of infection | Estimated during analysis | | |
| **$\tau$** | Treatment seeking rate (per day) | 0.97 | (0.84,0.99) | [19] |
| **$\delta$** | Natural recovery rate (per day) | 0.0035 | (0.0014, 0.017) | [20,22] |
| **r** | Recovery rate after antimalarial treatment (AL; per day) | 0.167 | (0.125, 0.25) | [20,23] |
| **$\rho$** | Loss of immunity (per day) | 0.0027 | (0.000055,0.011) | [18] |
| **$\theta$** | Relative infectiousness of asymptomatic infections | 0.467 | (0,0.50) | [17,24] |
| **$\zeta$** | Relative infectiousness of treated infections | 0.04 | (0.0,0.25) | [17] |
| **$\gamma_m$** | Rate of onset of infectiousness in mosquitoes (per day) | 0.1 | (0.07, 0.2) | [17,18] |
| **$\mu_m$** | Mosquito mortality rate (per day) | 0.033 | (0.0010,0.10) | [18] |

In this study, we assume that mosquito dynamics are static [17]. Consequently, they have a relatively rapid generation turnover and are highly responsive to changes in the proportion of infected humans. Hence, we simplify the vector equations and determine the number of humans who become infected under the prevailing model conditions by focusing on a single force of infection. That way, it allows us to run our simulation without considering the changes in vector dynamics. We derived the force of infection and model equations in the supplementary file 1. Furthermore, we introduced a seasonal forcing equation (described in the supplementary file) to mimic Zambia’s seasonal transmission pattern using rain data from the Climate Hazards Group InfraRed Precipitation with Station data.

### Reactive case detection rate

The rate of detecting cases τRCD was defined based on Njau and Silal et al. (2021) as equation 1, where *cov_RCD_* is the proportion of index cases that are followed up *incidence*, is the number of new index cases at the health centre, while *sample* is the number of people screened that are within the proximity of the index case [17]. Furthermore, the *pop* and *RDTsensitivity* are population in the model and the sensitivity of RDTs used during the intervention, respectively [17].

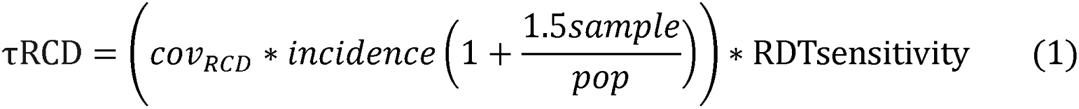

To incorporate CHWs, we define *cov_RCD_* as a function of CHWs and index cases as presented in equation 2, where we estimated the numerator as the average number of index cases investigated by a single CHW per day based on the information provided by Larsen et al. (2017) [4]. Here, 333 CHWs investigated approximately 854 index cases in one year in some low transmission areas of Zambia. Therefore, we divided the total number of index cases investigated by the number of CHWs and then by the number of days in a year, which gives us equation 2.

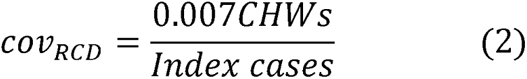

### Incorporating reactive Focal Mass Drug Administration

We additionally explore reactive fMDA as one of the measures to reduce malaria cases by interrupting transmission in the HFCA. Thus, all individuals within the proximity of the index case receives treatment, implying that, infections individuals with low levels of parasitaemia who would not have been detected by RDT gets cleared of parasite [16,22,24,25]. Similarly, the suspectable and exposed individuals are prevented from transitioning to the infectious category to transmit, thereby interrupting the transmission cycle with the HFCA [16,22,24,25]. Mathematical equations describing the incorporation are described by supplementary file equation 21.

### Reactive case detection improvement scenarios

Supplementary Table 1 presents a summary of simulated scenarios compared to the baseline. These scenarios were formulated to address common situations that often result in inefficient implementation of RCD. The purpose of formulating these scenarios was to assess their potential to achieve results similar to or better than the baseline. In these simulations, we assume that other interventions implemented to keep the low transmission status in the HFCA remain consistent and that situations only affected RCD implementation.

The baseline configuration assumed 20 community health workers per health centre dedicated to RCD, with each CHW representing a health post serving 500 individuals. This configuration was based on the estimated population of 10,000 in the HFCA, with an area coverage radius of 140 meters per index case [4,5,15]. The reaction time, which refers to the time taken to respond to reported cases, was set at three days. No fMDA was implemented, and RDTs had a sensitivity of 84% (Sup. Table 1).

In the “Increased number of CHWs” scenario, the number of CHWs per HFCA was increased to 30, while all other parameters remained unchanged from the baseline scenario (Table 2). This adjustment aimed to improve coverage and response capabilities within the same coverage radius (Sup. Table 1). In the Increased Radius (250 meters) scenario, the coverage area was expanded by increasing the radius to 250 meters, while keeping the number of community health workers and other parameters constant (Table 2). Similarly, in the “Increased radius (450 meters)” scenario, the coverage radius was further increased to 450 meters (Table 2).

**Table 2.**
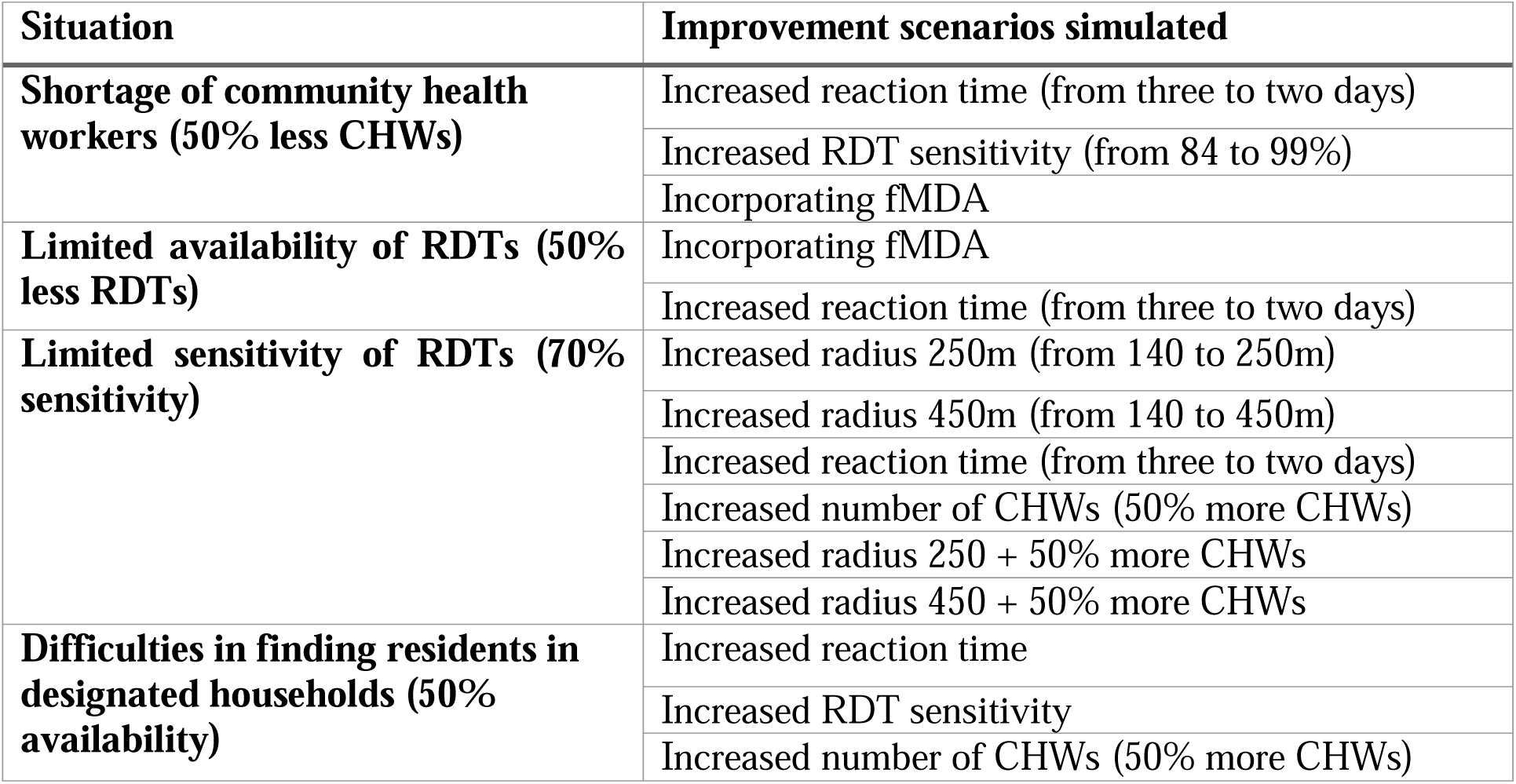
Situations and potential improvement scenarios.

Furthermore, we explored scenarios that combined an increase in the number of CHWs (30) with an increase in the coverage radius, both for 250 and 450 meters (Sup. Table 1). These scenarios aimed to improve both personnel and coverage area to improve intervention outcomes. Also, we simulated fMDA with the assumption that acceptance, coverage and drug efficacy all remain constant. The scenarios “Improved reaction time” (IRT) and “Increased RDT Sensitivity” had the same parameters as the baseline, except for changes in the reaction time (2 days) and RDT sensitivity (99%) (Sup. Table 1). The RDT sensitivity was set at this level, with the potential to be replaced by polymerase chain reaction as the testing option (Sup. Table 1).

### Challenges/situations affecting reactive case detection and potential improvement scenarios

Table 2 summarizes the common situations mentioned elsewhere that may lead to reduced efficacy of implementation of RCD. It also presents simulated improvement scenarios to assess their potential for maintaining or improving the effectiveness of RCD when faced with potential impediments. Here, we assumed that if the health facility is faced with a situation such as a shortage of CHWs, it is unable to immediately replenish them but requires conducting a different improvement measure that may maintain or improve the results of RCD.

### Malaria risk stratification

In Zambia, there is an annual program that stratifies each HFCA based on malaria transmission intensity levels. These levels are categorized as "no malaria" (level 0), "very low" (level 1, between 0 and 50 cases per 1000 population/year), "low" (level 2, between 50 and 200 cases per 1000 population/year), "moderate" (level 3, between 200 and 500 cases per 1000 population/year), and "high" (level 4, with over 500 cases per 1000 population/year [3,26]. In this study, we used the same stratification as thresholds to ascertain that our model outputs are within the malaria risk classifications and to inform the impact of RCD hurdles and their respective improvement measures while assuming that all other interventions remain implemented at a constant rate. Our model was run at a HFCA level (10,000 individuals) and day as the unit of change. Therefore, for the HFCA to qualify as a low transmission area (less 200 cases per year), it is required to have approximately less than 5.48 cases per day, thus, a sum of 2000 cases per 365 days. The total of 2,000 cases per 365 days is derived from each HFCA consisting of 10 populations of 1,000 people each, which, when multiplied by 200, results in 2,000.

### Uncertainty intervals for model results

To account for model uncertainty, we generated uncertainty intervals for all the results. We did this by running 100 simulations using randomly generated parameter values within the lower and upper bounds for all parameters in each model scenario. After generating results from 100 simulations for each scenario, we grouped daily malaria case values and obtained the median, 5th, and 95th quantile values for each day. The median value was used as the central value for each day, while the 5th and 95th quantile values were used as the lower and upper uncertainty values, respectively. This process was repeated for all model scenarios.

## Results

### Impact predictions for key challenges affecting the efficiency of reactive case detection

Figure 2 shows daily malaria cases for a hypothetical HFCA. In the figure, the baseline (*purple trend line*) represents a scenario in which all interventions including RCD are being implemented in accordance with recommended guidelines for low transmission areas, while the red horizontal dotted line represents the threshold at which the HFCA ceases to be classified as a low transmission area (Figure 2). Overall, simulating a 50% reduction in the literature-identified challenges affecting the effectiveness of RCD and reducing RDT sensitivity to 70% did not result in the HFCA completely exiting its low transmission status, indicated by the red dotted line (Figure 2). Nevertheless, a 50% reduction in the number of CHWs (*red*) and availability of RDTs (*blue*) for RCD resulted in the highest deviation of malaria cases from the baseline compared to the use of less sensitive RDTs (*light blue*) and not reaching 50% of individuals in their households (*green*), which had the least impact (Figure 2).

**Figure 2:**
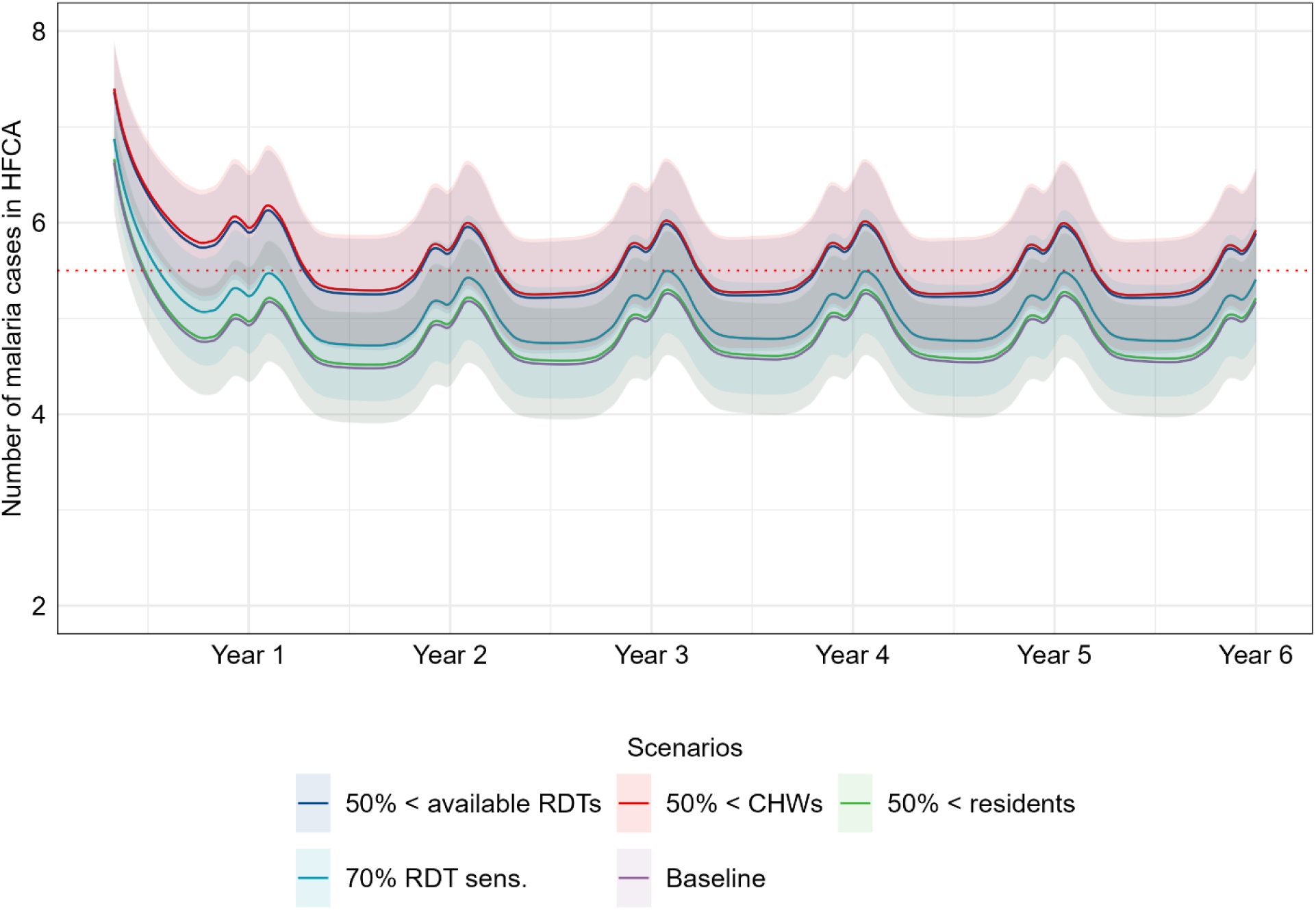
Impact prediction of key challenges impacting the efficiency of RCD. In this simulation, the four scenarios are compared to baseline and ability to influence malaria risk stratification status. In this figure, the red horizontal dotted line represents the threshold at which the HFCA remains a low-risk area. “50% < available RDTs” indicates a 50% reduction in the availability of RDTs, “50% < CHWs” represents a 50% shortage of CHWs, “50% < residents” denotes that only 50% of residents are available in designated households, and “70% RDT sens.” represents the use of RDTs with 70% sensitivity.

### Impact predictions of RCD improvement measures for shortage of community health workers

In the scenario of a 50% shortage of CHWs, as shown in Figures 2 and 3, results in the HFCA to near exiting the low-risk strata, with approximately 1914 cases per year (approximately 191.4 cases per 1000 population per year). The simulation results shown in Figure 3 demonstrate that incorporating fMDA (*light green*) as an improvement measure results in relatively fewer cases compared to all other measures, including the baseline (*purple*). Increasing RDT sensitivity (*red*), even up to 99%, as an improvement measure made the least difference compared to the 50% CHW shortage scenario (Figure 3). However, improving the reaction time (*light blue*) from the recommended three days to two days resulted in relatively fewer cases but not less or equal to the number of in the baseline scenario (Figure 3).

**Figure 3:**
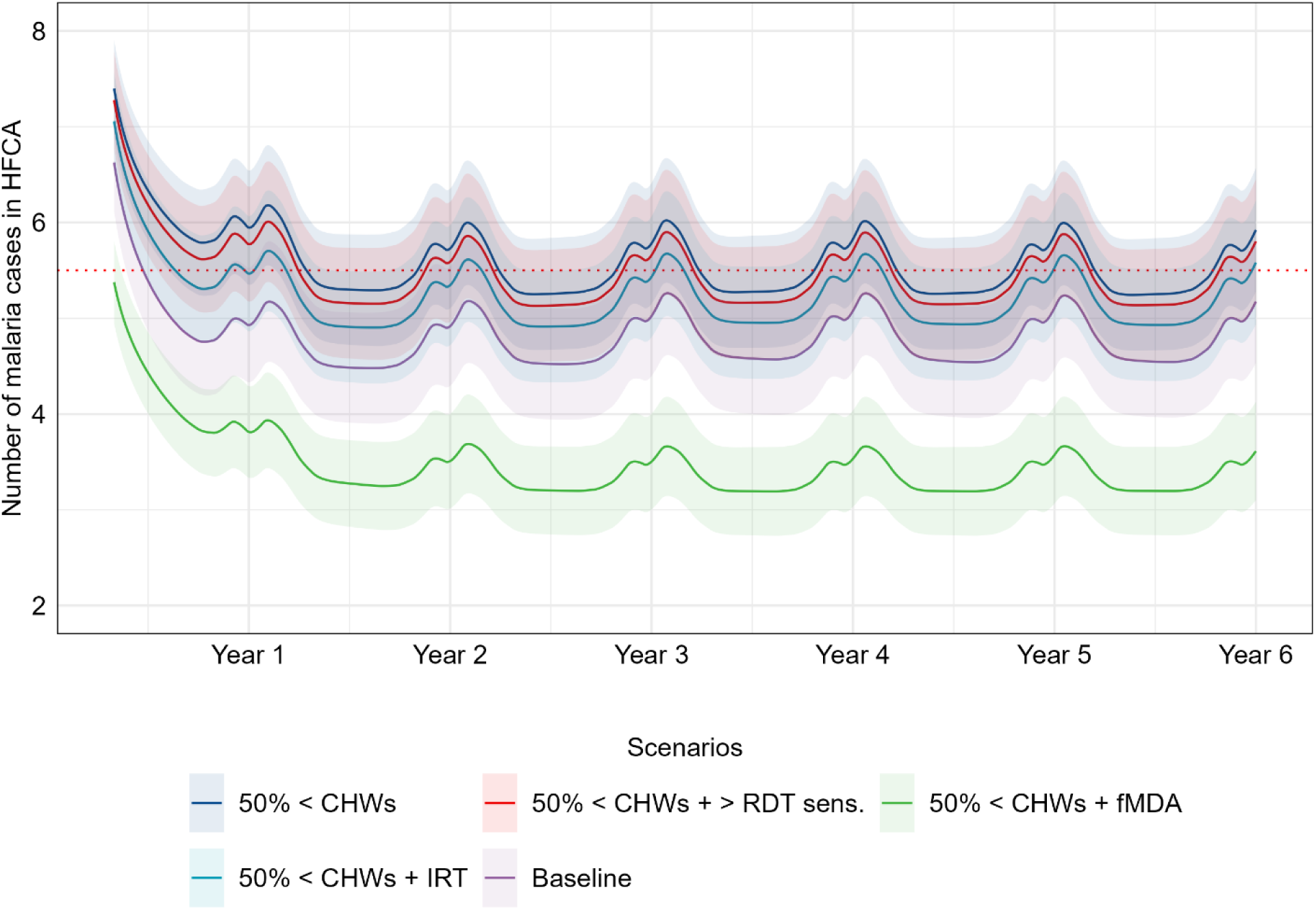
Impact predictions of improvement measures for shortage of CHWs. In this simulation, the number of CHWs was reduced by 50%, represented by “50% < CHWs,” i.e., from 20 to 10 CHWs per 10,000 population. The “50% < CHWs + RDT sens.” scenario depicts the use of more sensitive (99%) RDTs as an improvement measure to counter the impact of reduced CHWs. Similarly, “50% < CHWs + fMDA” and “50% < CHWs + IRT” represent using fMDA and improving the reaction time from three to two days, respectively as countermeasures.

### Impact predictions of RCD improvement measures for the limited availability and sensitivity of RDTs

Similar to the impact of a 50% shortage of CHWs, a 50% limited availability of RDTs, as depicted in Figures 2 and 4a, suggests that this scenario almost results in the HFCA exiting the low-risk strata, with approximately 1903 cases per year (approximately 190.3 cases per 1000 population per year).. The simulation results presented in Figure 4a indicate that the limited availability of RDTs (*blue*) is better improved by incorporating fMDA (*red*) rather than improving the reaction time (*green*) from three to two days.

**Figure 4:**
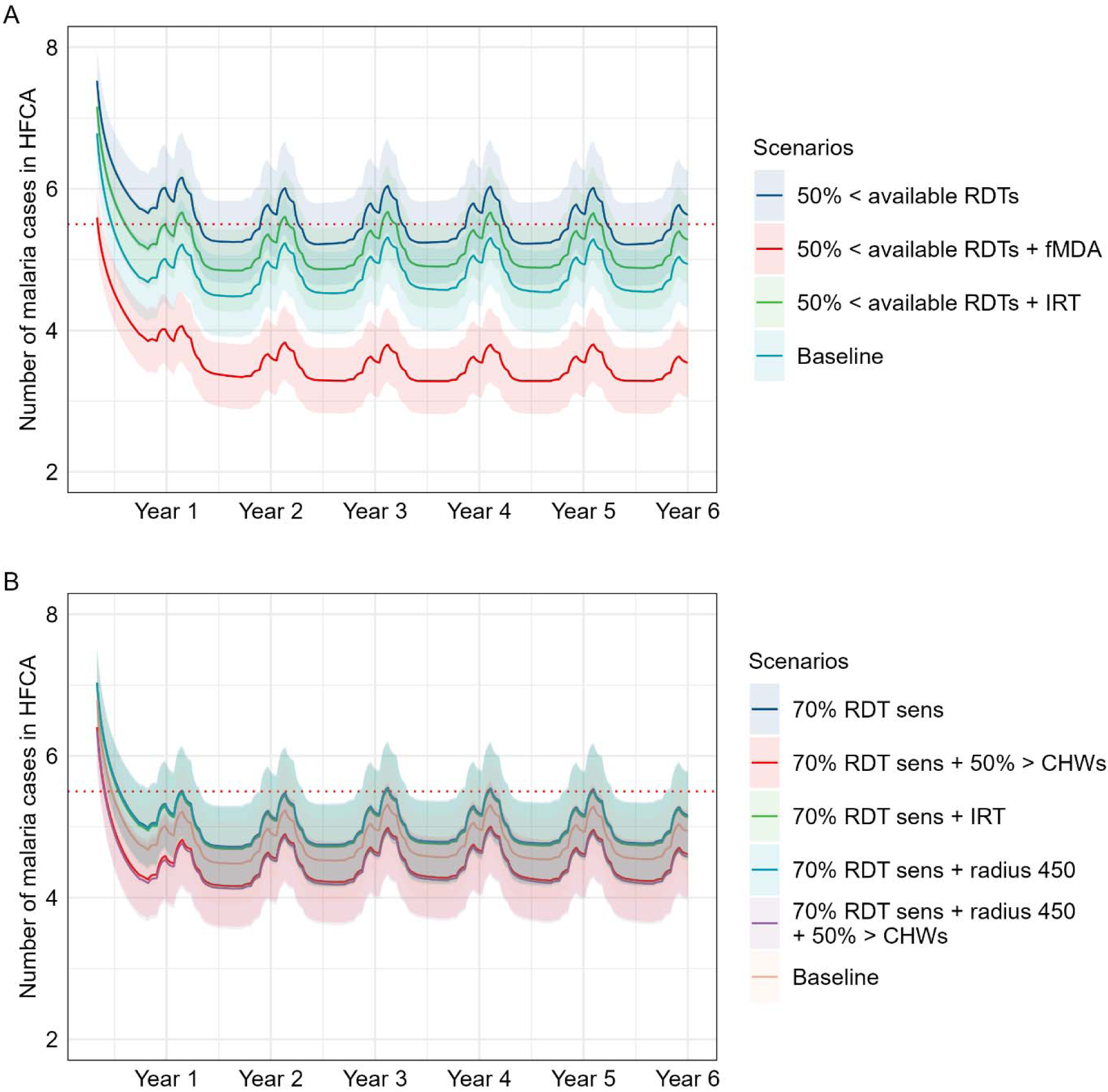
Impact predictions of RCD improvement measures for the limited availability and sensitivity of RDTs: (**A**) 50% of the secondary cases were tested, while fMDA (50% < available RDTs + fMDA) and a two-day reaction time (50% < available RDTs + IRT) were simulated as improvement measures. (**B**) Simulation of RDT sensitivity reduced to 70% and increase of radiu (70% RDT sens. + radius 450), reaction time (70% RDT sens. + IRT), and 50% more CHW (70% RDT sens. + 50% > CHWs) as improvement measures. Furthermore, in **B**, the blue, light blue and green share a similar trend, therefore obscuring each other, indicating negligible effect from the reduced RDT sensitivity.

On the other hand, the scenario of limited sensitivity of RDTs (*blue obscured by green*) is best improved when an increase in the number of CHWs dedicated to RCD and an expanded radius (*red*) as shown in Figure 4b. Thus, combining “increased the number of CHWs” by 50%, from 20 per HFCA to 30, and “expanding the radius to 450 meters” results in relatively fewer cases than the baseline (*orange*). However, augmenting the CHW workforce alone also leads to an approximately similar trend (Figure 4b). Notably, increasing the radius and reaction time independently had a negligible impact (Figure 4b), suggesting that the increase in CHWs is the main contributor to case reduction in the increased CHWs and radius “combined” scenario. Furthermore, uncertainty intervals for the baseline, reduction in RDT sensitivity and the improvement measures overlap signifying, potential for non-difference among (Figure 4b).

### Impact predictions of RCD improvement measures for difficulties in finding residents in designated households

Impact predictions of all investigated improvement measures for difficulties in finding residents in designated households resulted in relatively fewer cases than the baseline (*purple*) as depicted in Figure 5. The success of the improvement measure is attributed to the impact of not finding individuals in households (*blue almost sharing the same trend as purple baseline*) had negligible effect on the overall number of cases (Figure 5). Among the simulated improvement measures, improving the reaction time (*light blue*) from three to two days and increasing the number of CHWs by 50% (*green*), had the most impact at addressing the issue, as observed in Figure 5. Conversely, increasing RDT sensitivity (*red*), even up to 99%, had the least improvement but showed slightly lower infections than baseline (Figure 5). Similar to the RDT sensitivity reduction scenarios in Figure 4b, the difficulties in reaching residents scenarios show in Figure 5 the uncertainty intervals for the baseline and the improvement measures overlap signifying, potential for non-difference among them.

**Figure 5:**
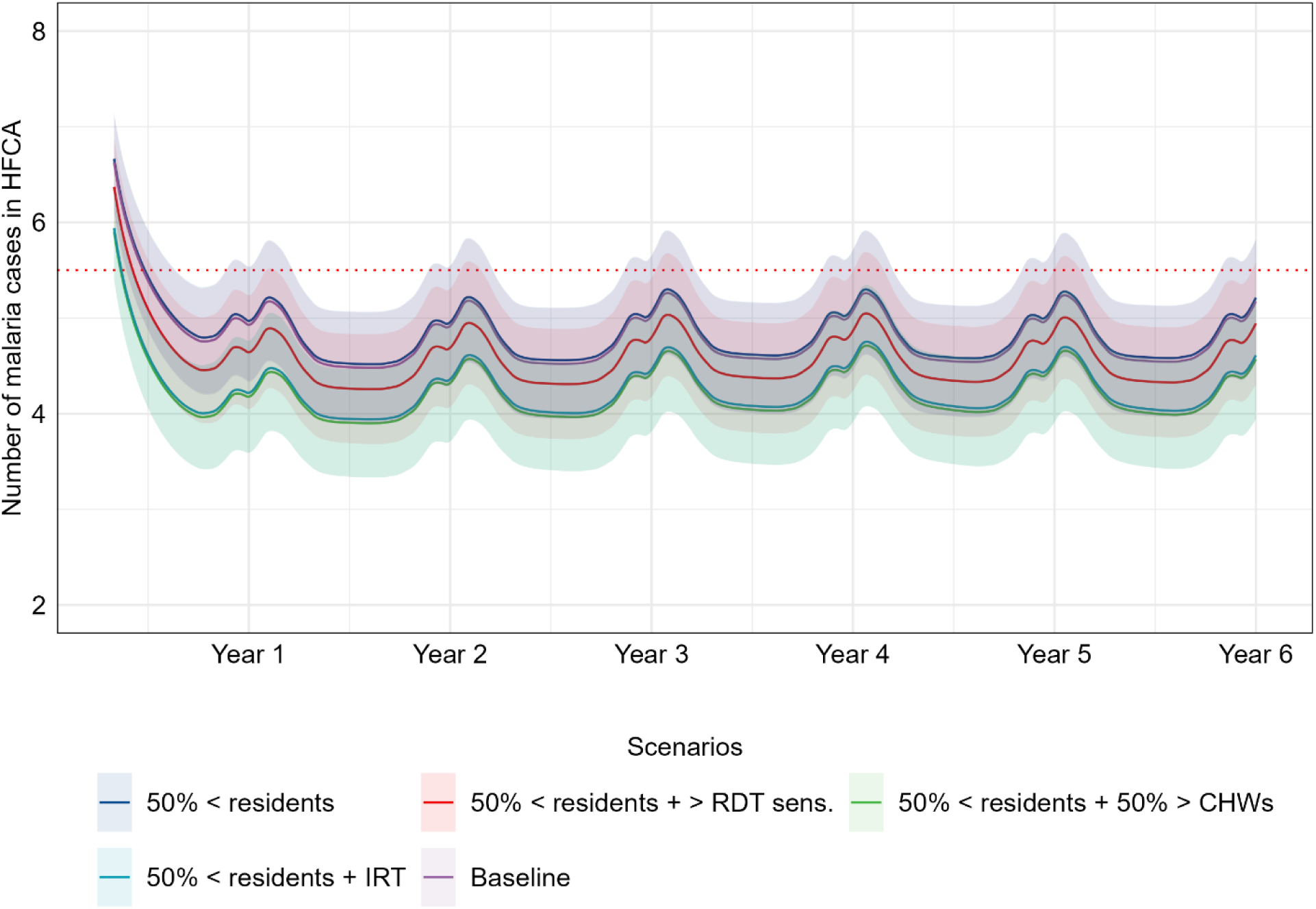
Impact predictions of improvement measures for difficulties in finding residents in designated households. In this simulation, residents’ availability was reduced by 50% (50% < residents), while a two-day reaction time (50% < residents + IRT), 99% RDT sensitivity (50% < residents + RDT sens.), and 50% CHWs increase (50% < residents + 50 > CHWs) were simulated separately as improvement measures.

## Discussion

We simulated an RCD focused model using parameters and data conforming to Zambia’s low transmission areas using a deterministic non-linear ordinary differential equation (ODE) model. Our primary objectives were to assess the impact of various literature-informed challenges affecting RCD to reduce malaria cases and their potential improvement measures. The analysis was undertaken with the purpose of informing the order for prioritizing the RCD challenges and guiding the appropriate improvement measures for each respective challenge. Considering that Zambia’s malaria risk stratification is done at a HFCA level and RCD is only conducted in low transmission HFCAs, the study was set up at this level.

The simulated impact of RCD challenges on malaria cases within a HFCA revealed that a shortage of CHWs and RDTs was predicted to have the most negative impact on RCD. In contrast, not finding individuals in households was predicted to have least impact. In scenarios where the availability of CHWs and RDTs was reduced by 50%, while keeping other parameters constant, annual malaria cases increased by approximately 17%. In both cases, the only effective countermeasure was the incorporation of fMDA, which resulted in an approximate 37% reduction in annual cases within the HFCA. However, using more sensitive RDTs and reducing the response time to counter the 50% shortage in CHWs resulted in only two and seven percent reductions in annual cases, respectively. Furthermore, in scenarios where RDT sensitivity was reduced to 70%, annual cases increased by 5%, while reducing the availability of individuals for testing by 50% led to approximately 0.8% increase in annual cases within the HFCA. In both scenarios, increasing the number of CHWs by 50% to offset their negative effects on RCD resulted in approximately 12% and 14% decreases in annual cases when using 70% sensitive RDTs and when availability of individuals was reduced by 50%, respectively. Additionally, in both scenarios, increasing CHWs by 50% as a countermeasure led to relatively fewer cases compared to adjusting the reaction time from three to two days or increasing the screening radius from an index case up to 450m from the initial 140m. However, combining an increase in radius with a 50% increase in CHWs only reduced the number of annual cases by 13%, compared to the 12% reduction observed when only CHWs were increased to counter a 70% reduction in RDT sensitivity.

Considering that RCD operations in settings like Zambia are primarily conducted by CHWs, the number of index cases investigated directly depends on the number of CHWs, as set up in our model[4,27]. Our findings suggest that having more CHWs available for RCD results in more index case follow-ups. As such, our finding that the number of CHWs has the most substantial effect on the effectiveness of RCD aligns with Chitnis et al. (2019), who suggested that RCD is only successful in low transmission areas if many index cases are followed up [9]. However, Reiker et al. (2019) added that it is important to assess follow-up capacity rather than merely considering the actual number of cases. They argued that the potential number of index cases is limited by those who either do not seek official care or are asymptomatic, suggesting that the number of investigated index cases should be adjusted based on treatment-seeking behaviour [9,10].

Consistent with several studies, our research also indicates that MDA interventions may be the most effective alternative in various situations where RCD’s effectiveness is limited. For instance, Ntunku et al. (2022) noted that RCD requires notably more personnel time compared to fMDA and therefore uses fewer resources [28]. Additionally, even though we modelled our baseline scenario with assumption that RCD was conducted perfectly, it did not result in zero infections in the HFCA over time. This supports findings from other studies that highlight that the ability of RCD to eliminate malaria depends on multiple factors, such as environmental risks and other archetypical factors, which our model may not have considered [5,9,10,14]. Nevertheless, our model results demonstrated that RCD managed to maintain the number of cases within the low transmission strata, and when we reduced the number of CHWs by half, the number of cases nearly surpassed the low transmission threshold.

The study offers valuable insights into the challenges that impact the effectiveness of RCD and potential countermeasures. Implementers can utilize these insights to evaluate their resource capacity and combinations to suit an ideal RCD programme. For example, if the ratio of CHWs dedicated to RCD to the catchment population exceeds 1:1000 or if the health centre frequently experiences stockouts of RDTs, it might be necessary to consider fMDA as an alternative measure to interrupt infections in the HFCA. Otherwise, other improvements are likely to be ineffective. In situations where only less sensitive RDTs are available, the health facility may consider recruiting more CHWs to increase the number of index cases followed up as an improvement measure. However, failing to find secondary individuals should have relatively less priority compared to addressing other RCD challenges. In Zambia, the COVID-19 pandemic period poses a good example where the shortage of RDTs scenario is more applicable. In the same period, the Zambian healthcare system had 16,000 CHWs trained to undertake community-focused malaria interventions, but a number of them were inactive [3]. Reasons for the inactivity included the limited supply of antimalarial drugs (ACTs) and RDTs caused by COVID-19-induced supply chain interruptions [3].

It is worth noting that, the context governing the setup of how RCD is conducted may vary the outcomes of the results presented in this study. Furthermore, it is essential to note that this study faces certain limitations, including the assumption that CHWs possess “perfect” knowledge of how to conduct RCD. This is contrary to the evaluation by Searle et al. (2016), where it was established that the operational challenges for inadequate implementation of RCD may have also been due to various CHWs’ related inadequate technical capacity, such as the inability to distinguish the houses within the prescribed radius from the index case’s house [5,29]. Additionally, our model assumed that RCD was conducted daily, which is somewhat unrealistic given that CHWs have roles other than conducting RCD in the HFCA. Also, certain operational qualifiers/disqualifiers for RCD/MDA, such as travel history, season, and risk for drug resistance, were ignored. Furthermore, the study assumed that antimalarial drugs were in abundant supply despite the shortage of RDTs, which may be unlikely, as alluded to earlier that both were in short supply during the peak of COVID-19 [3]. As such, our model may be overstating the number of secondary cases that may be treated via MDA if RDTs are in short supply. Moreover, the model was not calibrated to any real data; therefore, all findings remain hypothetical and do not necessarily represent any true HFCA. Nevertheless, the outcome trends for the generated scenarios and the approach may be extended to other low-transmission catchment areas of Zambia with similar characteristics.

Based on results from [11,12,14–16,25,29–34] and this study, it is evident that the success of an RCD program in a Zambia-like setup highly depends on CHWs. However, their involvement is voluntary and primarily influenced by non-formal incentives, often provided by donors [29,35].

Finding sustainable means such as following World Health Organization’s 2018 guidelines for CHWs remuneration to ensure the sufficient availability of CHWs may guarantee continued RCD contributions towards maintaining stable malaria prevalence and ultimately contributing to achieving elimination [35]. The current CHWs’ contribution towards malaria programming and incentive situation warrants detailed research to optimize RCD implementation while considering potentially sustainable motivating incentives and monitoring and evaluation to maintain an optimal RCD programme. Furthermore, the coming of COVID-19, which interrupted several supply chain mechanisms including that for malaria supplies is a wake-up call to further look into improvement measures for other interventions other than RCD. The interventions of interest may include interruption of case management (antimalarial supplies and test kits), bednets, and IRS insecticides.

Lastly, considering the complexity of malaria transmission and several assumptions that were imposed on this study’s model. To make the modelling results more applicable across different contexts, we have research plans to stratify regions such as HFCAs into transmission archetypes based on malaria risk profiles, ecological, environmental, health system, and socioeconomic factors, and tailoring interventions to suit specific archetypes may improve the outcomes and expedite the contribution to malaria elimination. The archetyping approach may guide which low transmission areas will benefit most from RCD instead of alternative interventions or a combination of interventions. Conversely, interventions requiring fewer resources than RCD to avert the same number of malaria cases in certain archetypes may be opted for in place of RCD or a combination of interventions. Furthermore, the archetype-based targeting approach can also inform the optimization of interventions, including RCD. This way, the potential resources required to achieve elimination can be identified and planned for effectively.

## Conclusion

This study contributes to the existing research literature by examining the impact of challenges faced by RCD on malaria cases at an HFCA level. It also highlights the effectiveness of potential improvement measures for these challenges. The study used mathematical modelling to simulate several scenarios to mimic RCD challenges and their respective potential improvement measures in a Southern province of Zambia-like setting.

The exploration of improvement measures for RCD provided in this study offers opportunities to focus on measures that yield relatively better outcomes in specific situations. This approach may enhance the effectiveness of RCD in resource-constrained settings. Consequently, the contribution of RCD to malaria elimination may remain substantial, despite any challenges that may arise during its implementation.

## Data Availability

All data produced in the present work are contained in the manuscript

## Acknowledgements

The authors acknowledge the National Malaria Control Centre and the Modelling and Simulation Hub, Africa team for the contribution to ideas and feedback and code review.

## Author Contributions

Conceptualization, C.C., S.S.; Methodology, C.C and S.S; Software, C.C.; Formal analysis, C.C.; Investigation, C.C.; Resources, SS.; Writing—original draft, C.C.; Writing—review & editing, S.S.; Visualization, C.C.; Supervision, S.S.; Funding acquisition, S.S. All authors have read and agreed to the published version of the manuscript.

## Institutional Review Board Statement

Not applicable.

## Informed Consent Statement

Not applicable.

## Data Availability Statement

Not applicable.

## Supporting Information

**S1 File: Supplementary appendix**

