## supplementary file 1 for "Exploring measures to increase detection of malaria cases through reactive case detection in a Southern Province of Zambia-like Setup: A modelling study"

Chilochibi Chiziba[1*], Busiku Hamainza[2], Japhet Chiwaula[2], Sheetal Silal[1,3]

[1] Modelling and Simulation Hub, Africa, Department of Statistical Sciences, University of Cape Town, Cape Town, South Africa

[2]Ministry of Health, National Malaria Control Centre, Lusaka, Zambia

[3]Nuffield Department of Medicine, Centre for Global Health, University of Oxford, Oxford, United Kingdom

### Force of Infection definition

In this study, we assume that mosquito dynamics are static [1]. Consequently, they have a relatively rapid generation turnover and are highly responsive to changes in the proportion of infected humans [1]. Hence, we simplify the vector equations and determine the number of humans who become infected under the prevailing model conditions by focusing on a single force of infection [1]. That way, it allows us to run our simulation without considering the changes in the vector dynamics [1]. Mathematically, the model takes a simple SEIR-SEI model, where the I in the human model is disaggregated into the clinical C, asymptomatic A, and treated X and V populations. As such, we may reaggregate for easy derivation of the force of infection. Thus, $I = C + \theta A + \zeta X + \zeta V$. Therefore, we define the human population P as $P = S + E + I + R$, while the vector population is defined as M = $S_m + E_m + I_m$. As such, we define the model as follows.

$$\frac{dS}{dt} = \mu_h P - a\frac{M}{P}b\frac{I_m}{M}S + \rho R - \mu_h S \quad (1)$$

$$\frac{dE}{dt} = a\frac{M}{P}b\frac{I_m}{M}S - (\gamma_h + \mu_h)E \quad (2)$$

$$\frac{dI}{dt} = \gamma_h E - (r + \mu_h)I \quad (3)$$

$$\frac{dI}{dt} = rI - (\rho + \mu_h)R \quad (4)$$

$$\frac{dS_m}{dt} = \mu_m M - ac\frac{I}{P}S_m - \mu_m S_m \qquad (5)$$

$$\frac{dE_m}{dt} = ac\frac{I}{P}S_m - (\gamma_m + \mu_m)E_m \qquad (6)$$

$$\frac{dI_m}{dt} = \gamma_m E_m - \mu_m I_m \qquad (7)$$

In this study, we assume that mosquitoes are in a state of equilibrium (no change in mosquito population). Consequently, they have a relatively rapid generation turnover and are highly responsive to changes in the proportion of infected humans. Hence, we simplify the vector equations and determine the number of humans who become infected under the prevailing model conditions by focusing on a single force of infection as follows. That way, it allows us to run our simulation without considering the changes in the vector dynamics.

Firstly, we obtain the vector and human force of infections from equations 2 and 6 and define them as equations 8 and 9, respectively.

$$\lambda_m = ac\frac{I}{P} \qquad (8)$$

$$\lambda_h = a\frac{M}{P}b\frac{I_m}{M} = ab\frac{I_m}{P} \qquad (9)$$

We proceed by making $I$ the subject of the formula by setting vector equations 6 and 7 equal to 0, which become equations 10 and 11.

$$E_m = \frac{\mu_m}{\lambda_m}I_m \qquad (10)$$

$$ac\frac{I}{P}S_m = (\gamma_m + \mu_m)E_m \qquad (11)$$

As noted earlier that the mosquito population defined as $M = S_m + E_m + I_m$, therefore, if we substitute $\lambda_m = ac\frac{I}{P}$ into equations 10 and 11, we get equation 12, which can further be factorized into equations 13, 14, to 15.

$$\lambda_m\left(M - \frac{\mu_m}{\gamma_m}I_m - I_m\right) = (\gamma_m + \mu_m)\frac{\mu_m}{\gamma_m}I_m \qquad (12)$$

$$\lambda_m M - \lambda_m I\left(\frac{\lambda_m - \mu_m}{\gamma_m}\right) = \left(\frac{\gamma_m + \mu_m}{\gamma_m}\right)\mu_m I_m \qquad (13)$$

$$\lambda_m M = I_m \left(\frac{\gamma_m + \mu_m}{\gamma_m}\right)(\mu_m + \lambda_m) \quad (14)$$

$$I_m = \frac{\lambda_m M}{\frac{\gamma_m + \mu_m}{\gamma_m}(\mu_m + \lambda_m)} \quad (15)$$

We then substitute $I_m$ from equation 15 into the human force of infection $\lambda_m = ab\frac{I_m}{P}$ to get equation 16, which we further factorize into equation 17, after, which we assume that $m = \frac{M}{P}$ to get equation 18 as the force of infection.

$$= \frac{ab}{P} \frac{\lambda_m M}{\frac{\gamma_m + \mu_m}{\gamma_m}(\mu_m + \lambda_m)} \quad (16)$$

$$= \frac{ab\frac{M}{P}ac\frac{I}{P}}{\frac{\gamma_m + \mu_m}{\gamma_m}(\mu_m + ac\frac{I}{P})} \quad (17)$$

Therefore,

$$\lambda = \frac{a^2bcm\frac{I}{P}}{(\mu_m + ac\frac{I}{P})}\left(\frac{\gamma_m}{\gamma_m + \mu_m}\right) \quad (18)$$

In this study the mosquito-human ratio ***m*** was obtained by dividing the mean number of mosquitos per household (2.25 mosquitos) from the PMI Vectorlink Project Zambia Project Annual Entomology Report August 2021-July 2022 by the average Zambian size in Southern Province (4.9) [2]. We used the Southern province's values because it has a majority low transmission HFCAs [3].

Furthermore, we introduce a seasonality equation (equation 19) adapted from Njau and Silal et al. (2021) to the force to mimic Zambia's seasonal transmission pattern using rain data from the Climate Hazards Group Infrared Precipitation with Station data [1]. To achieve, data extraction, we downloaded monthly raster version of rainfall data for the year 2019. We chose 2019 rainfall data because, the parameters used in this study were obtained from studies from the same period. Furthermore, we downloaded the Zambia shape file from Humanitarian Data Exchange, which we overlayed on the raster files in RStudio using the terra package to extract monthly mean rainfall reading for the entire country. We opted to obtain the country level mean values because, low transmission HFCA are heterogeneously spread across the country. After extracting, the monthly data, we replicated it over a period of six years to align with the model period. Supplementary Figure 1A shows the resulting rainfall trend. In the figure, it is evident that rainfall picks around April, which aligns with malaria trends described in the malaria indicator survey.

$$sea(t) = 1 + rain * a * \cos(2\pi(t - \varphi)) \qquad (19)$$

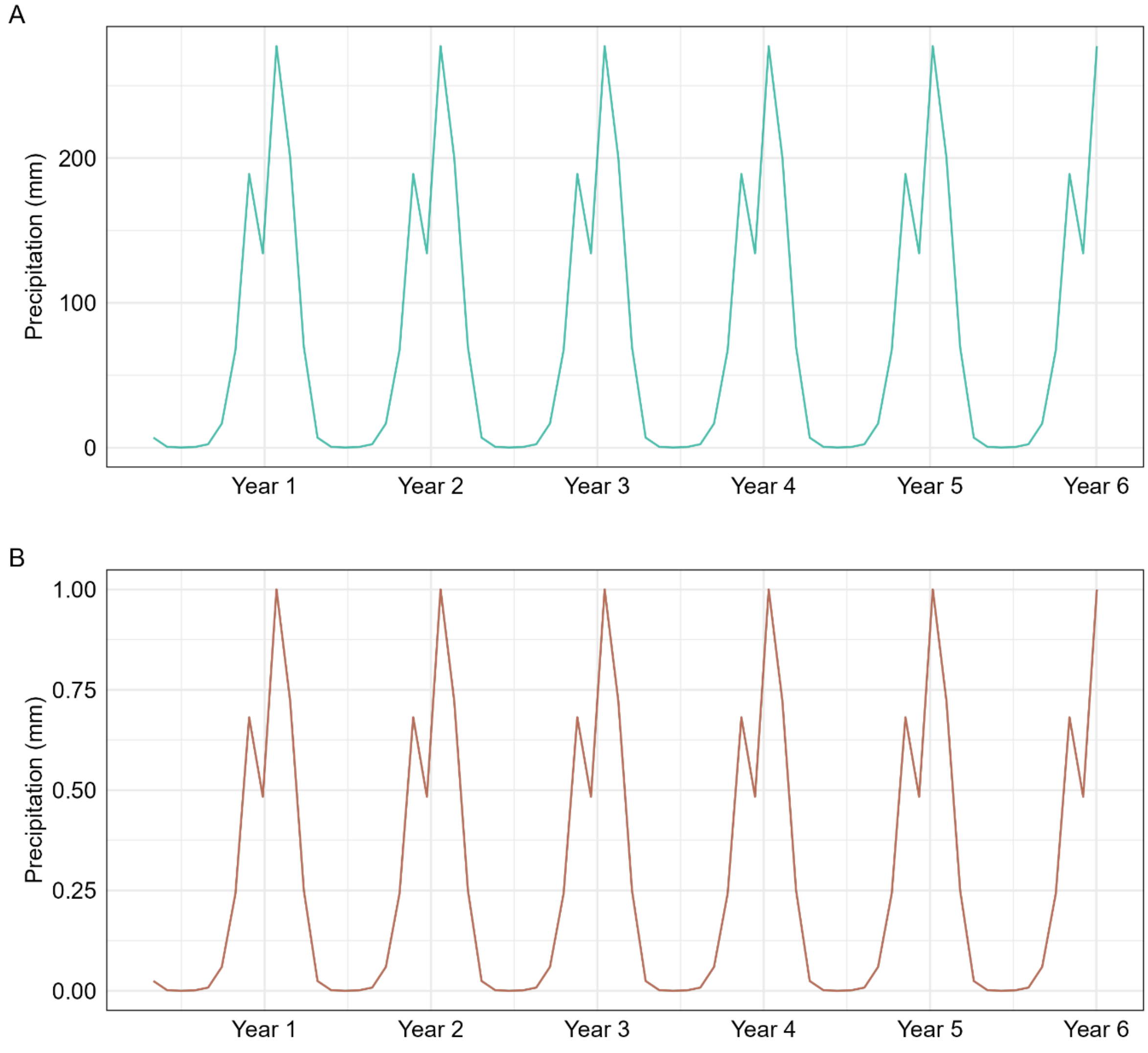


*Table 1: Precipitation trend in Zambia defined estimated from rain gauge and satellite observations. 3A shows the mean values in millimeters for the country, while 3B shows the standardized version of 3A.*

In equation 19, $\pi$ represents the peak month for transmission, rain is the monthly rain value, a is the amplitude for the seasonal variation, and *t* is the time step. The rain in the equation was standardized between zero and one (Supplementary Figure 1B) by subtracting the minimum monthly value of the year to each observation and dividing the difference by the difference between the max and min values for each year. Consequently, the seasonality function 19 was applied to the force of infection, making it equation 20 before adding the fMDA intervention. Considering that the *ode* time steps operate continuously, we used the *approx* function from the *stats* package to interpolate linear values between each for each time step t of the month, which represents days in our model.

$$\lambda = (sea(t)) \frac{a^2 bcm \frac{I}{P}}{(\mu_m + ac\frac{I}{P})} \left(\frac{\gamma_m}{\gamma_m + \mu_m}\right) \quad (20)$$

***Incorporating reactive Focal Mass Drug Administration***

We incorporated reactive fMDA as one of the measures to reduce malaria cases by interrupting transmission in the HFCA. Thus, all individuals within the proximity of the index case receive treatment, implying that, infectious individuals with low levels of parasitaemia who RDT would not have detected get cleared of parasite [4–6]. Similarly, the susceptive and exposed are prevented from transitioning to the infectious category to transmit, interrupting the transmission cycle with the HFCA [4–6] We incorporated fMDA by manipulating the force of infection with effectiveness, coverage, and acceptance from an MDA/fMDA trial in the southern Province of Zambia by Eisele et al. (2020). Where, 87.2\% effectiveness using dihydroartemisinin–piperaquine drug was established after attaining a 63–79\% household coverage with 87\% to 94\% community acceptance [7]. We assumed that incorporating fMDA in our model would yield similar impact from the trial, therefore, in each scenario where fMDA was considered, force of infection assumed equation 21.

$$\lambda = (1 - \text{fMDA}) * (sea(t)) \frac{a^2 bcm \frac{I}{P}}{(\mu_m + ac\frac{I}{P})} \left(\frac{\gamma_m}{\gamma_m + \mu_m}\right) \quad (21)$$

Where fMDA = effectiveness of fMDA * coverage * community acceptance of fMDA.

As such we mathematically define the RCD model as in figure 1 as follows.

$$\frac{dS}{dt} = -\lambda S + \rho \text{R} \quad (22)$$

$$\frac{dE}{dt} = \lambda S - pa\gamma \text{E} - (1 - \text{pa})\gamma \text{E} \quad (23)$$

$$\frac{dA}{dt} = pa\gamma \text{E} - (1 - \text{da})\delta A - da(\tau \text{RCD}) \quad (24)$$

$$\frac{dC}{dt} = (1 - pa)\gamma E - \tau RCD(1 - ga)C - ga(\tau)C \qquad (25)$$

$$\frac{dX}{dt} = ga(\tau)C - r(X) \qquad (26)$$

$$\frac{dV}{dt} = da(\tau RCD)A + \tau RCD(1 - da)C - r(V) \qquad (27)$$

$$\frac{dX}{dt} = (1 - da(\delta)A + r(X) + r(V) - \rho(R) \qquad (28)$$

**Simulation scenarios**

Supplementary Table 1 presents a comparison summary of various simulated scenarios against a baseline scenario, designed to address common inefficiencies in the implementation of Reactive Case Detection (RCD). The simulations assume consistent implementation of other interventions maintaining low malaria transmission, focusing solely on the effects on RCD implementation.

*Table 2: Simulation scenarios*

| Scenario | CHWs per Health center | Radius | Reaction time | fMDA | RDTS sensitivity |
|---|---|---|---|---|---|
| **Baseline** | 20 | 140 | 3 days | None | 0.84 |
| **Increased number of CHWs per Health center** | 30 | 140 | 3 days | None | 0.84 |
| **Increased radius 250** | 10 | 250m | 3 days | None | 0.84 |
| **Increased radius 450** | 10 | 450m | 3 days | None | 0.84 |
| **Increased radius 250 + Increased number of CHWs** | 30 | 250 | 3 days | None | 0.84 |
| **Increased radius 450 + Increased number of CHWs** | 30 | 450 | 3 days | None | 0.84 |
| **Increased reaction time** | 20 | 140 | 2 days | None | 0.84 |
| **Incorporating fMDA** | 20 | 140 | NA | Yes | NA |
| **Increased RDT sensitivity** | 20 | 140 | 3 days | None | 99% |

### Model parameter verification and sensitivity analysis results

*Table 3: Non-intervention parameter verification table showing the mean values for each compartment when the baseline parameter value was varied by the test value.*

| Symbol | Definition | Verification status |
|---|---|---|
| **a** | Human feeding rate per mosquito (per day) | Checked |
| **b** | Transmission efficiency from mosquito to human (per day) | Checked |
| **c** | Transmission efficiency human to mosquito (per day) | Checked |
| **pa** | Proportion of asymptomatic infections | Checked |
| **da** | Proportion of asymptomatic infections that get screened and treated through RCD | Checked |
| **ga** | Proportion of clinical infections that get treatment at health facility | Checked |
| $\gamma$ | *Rate of onset of infectiousness in humans (Incubation rate;* per day) | Checked |
| $\tau$ | Treatment seeking rate (per day) | Checked |
| $\delta$ | Natural recovery rare (per day) | Checked |
| $r$ | *Recovery rate after antimalarial treatment (AL;* per day) | Checked |
| $\rho$ | *Loss of immunity (*per day) | Checked |
| $\theta$ | *Relative infectiousness of asymptomatic infections* | Checked |
| $\zeta$ | *Relative infectiousness of treated infections* | Checked |
| $\gamma_m$ | *Rate of onset of infectiousness in mosquitoes* (per day) | Checked |
| $\mu_m$ | *Mosquito mortality rate* (per day) | Checked |

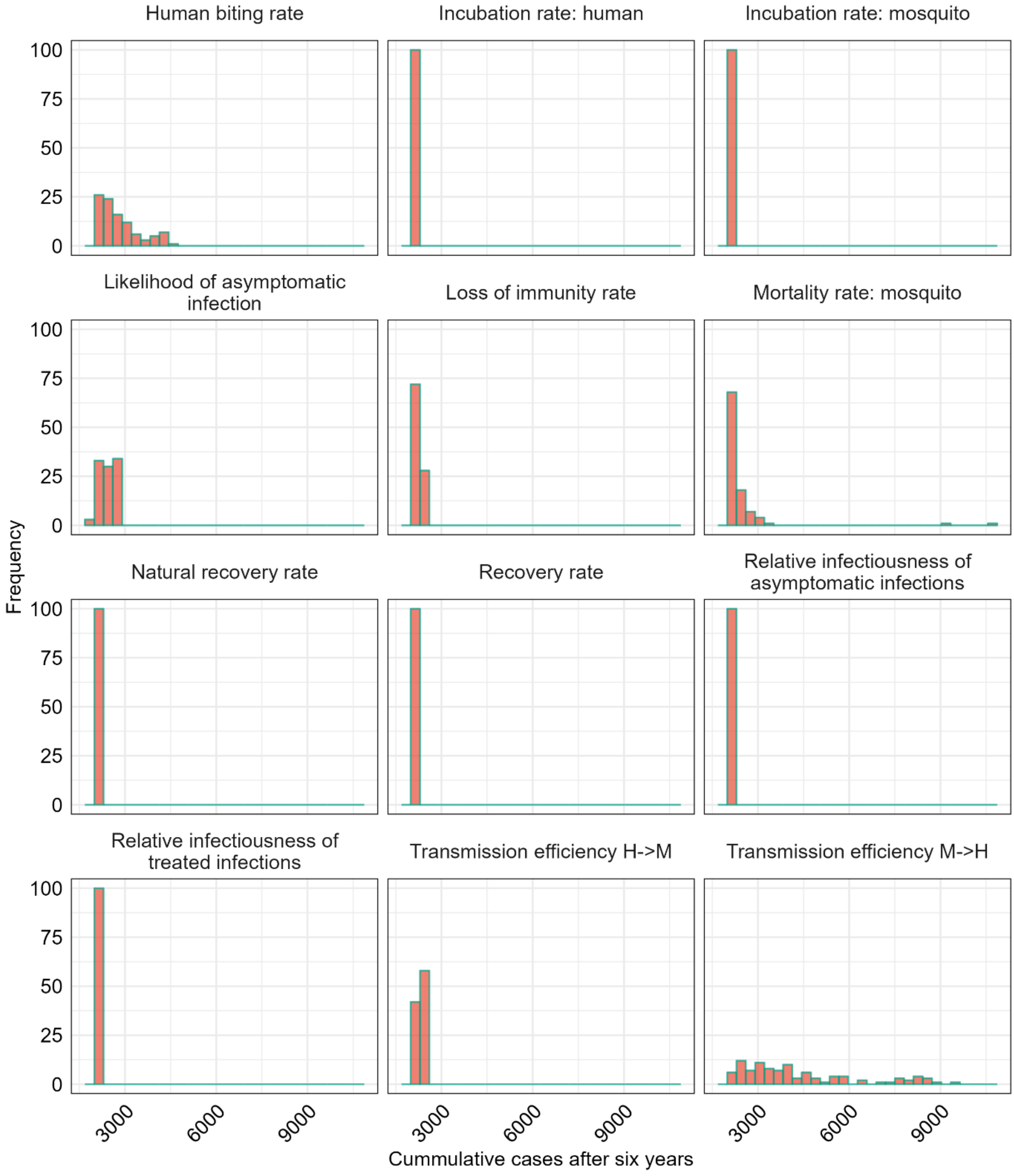


*Figure 1: Cumulative malaria cases observed after varying non-intervention related parameters. The parameter ranges considered are highlighted in manuscript Table 1.*

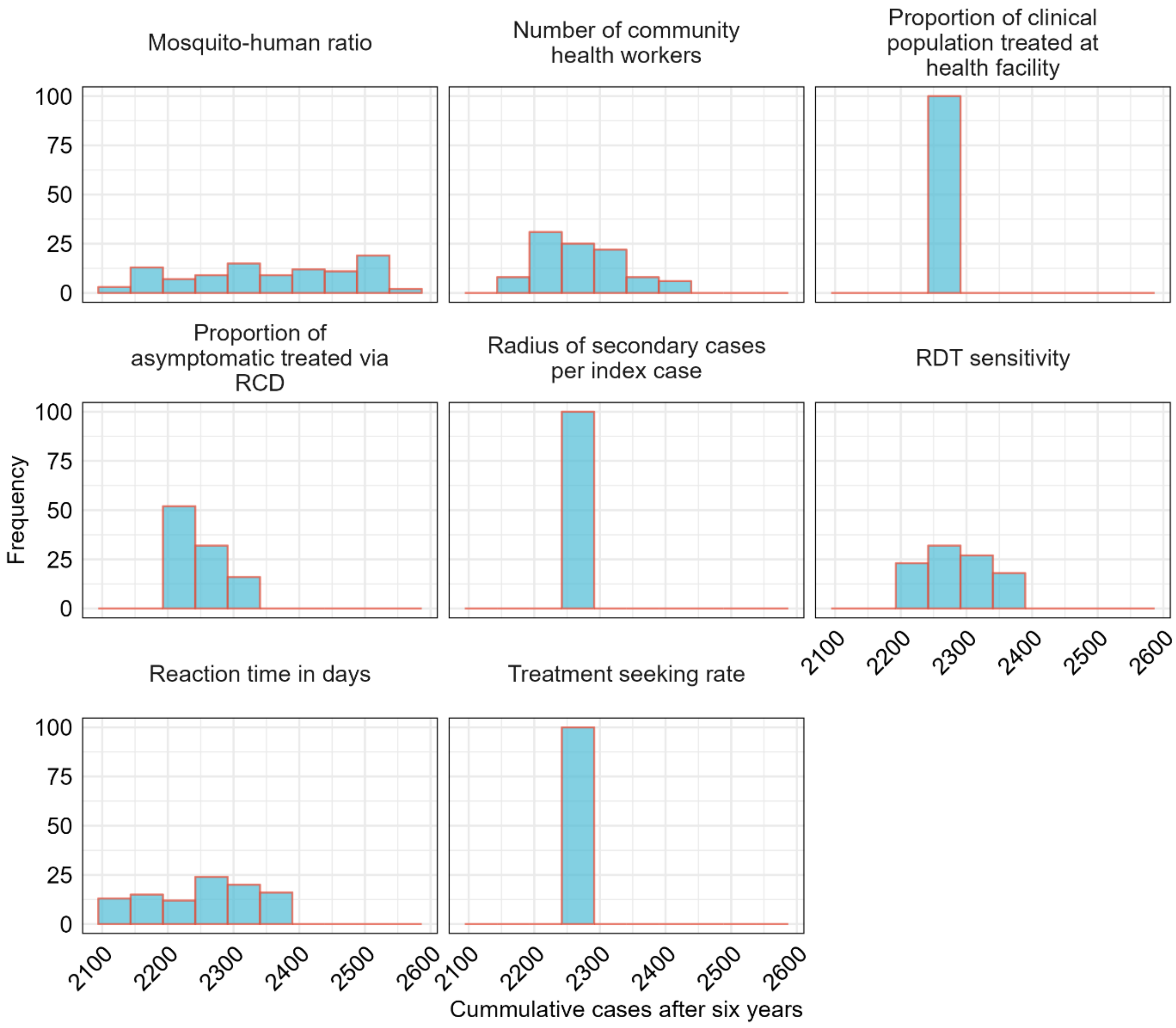


*Figure 2: Cumulative malaria cases observed after varying intervention related parameters. The parameter ranges considered are highlighted in manuscript 1.*

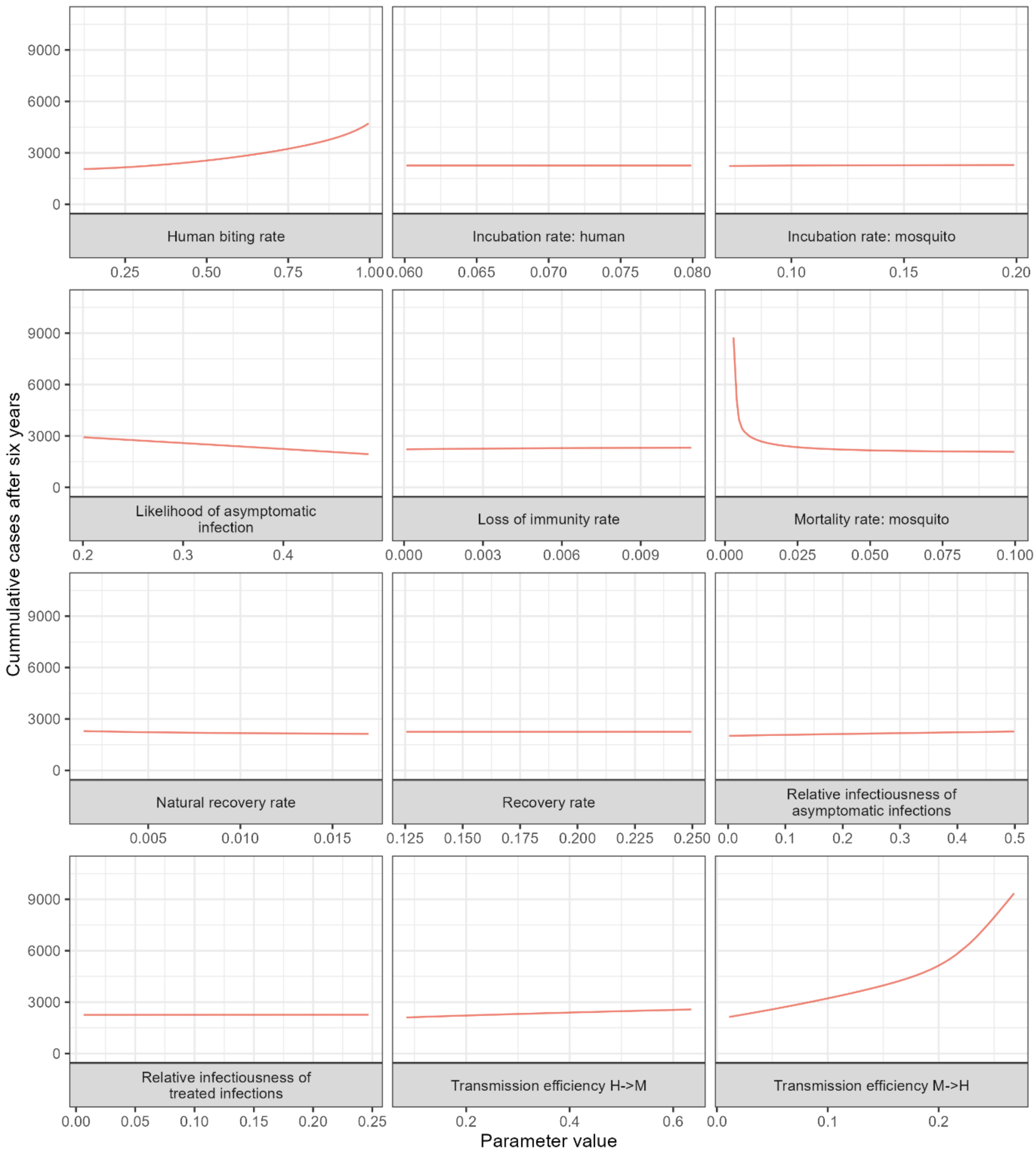


*Figure 3: Relationship between cumulative malaria cases observed after varying non-intervention related parameters and random values between their ranges. The parameter ranges considered are highlighted in manuscript 1.*

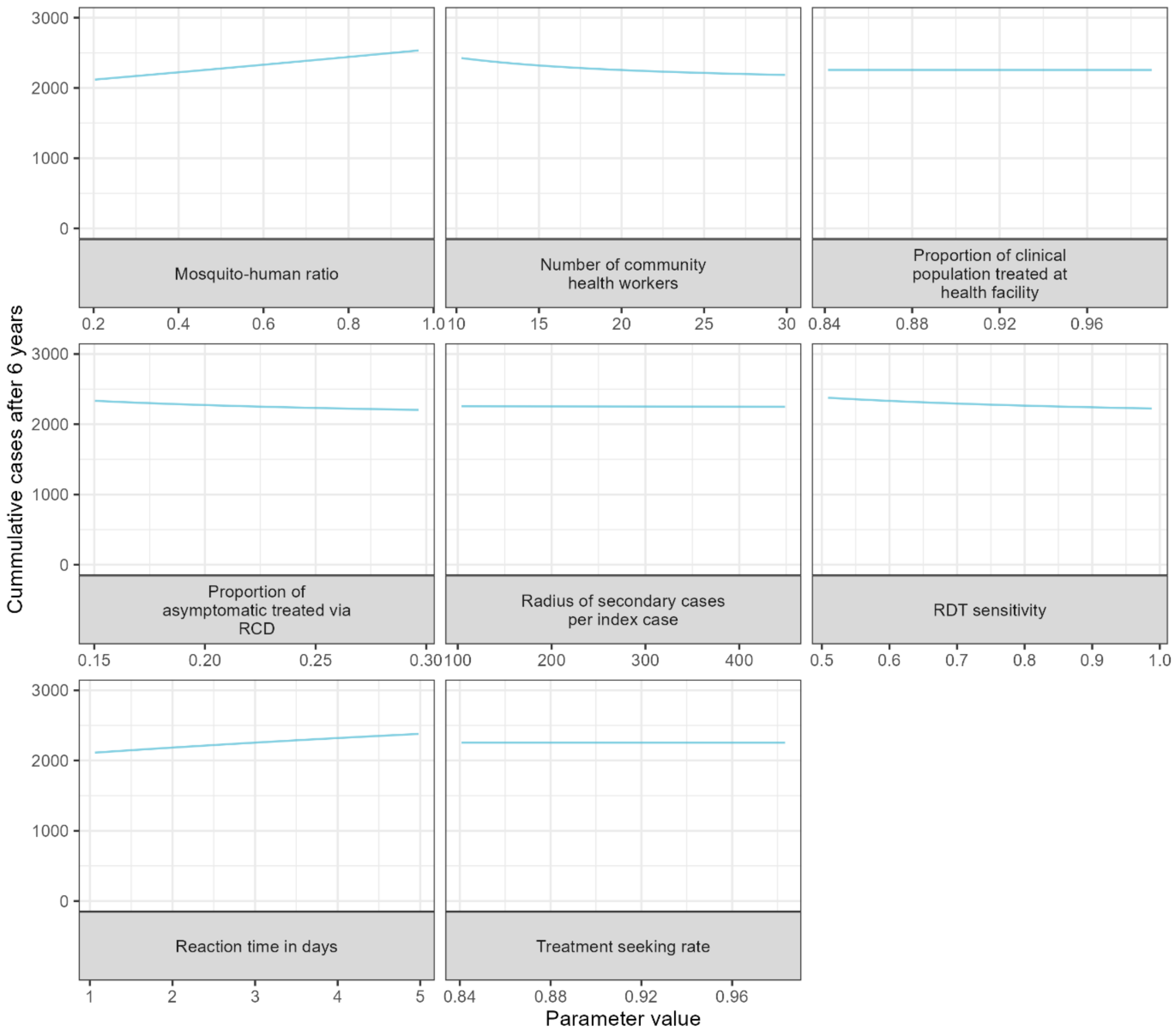


*Figure 4: Relationship between cumulative malaria cases observed after varying intervention related parameters and random values between their ranges. The parameter ranges considered are highlighted in manuscript 1.*

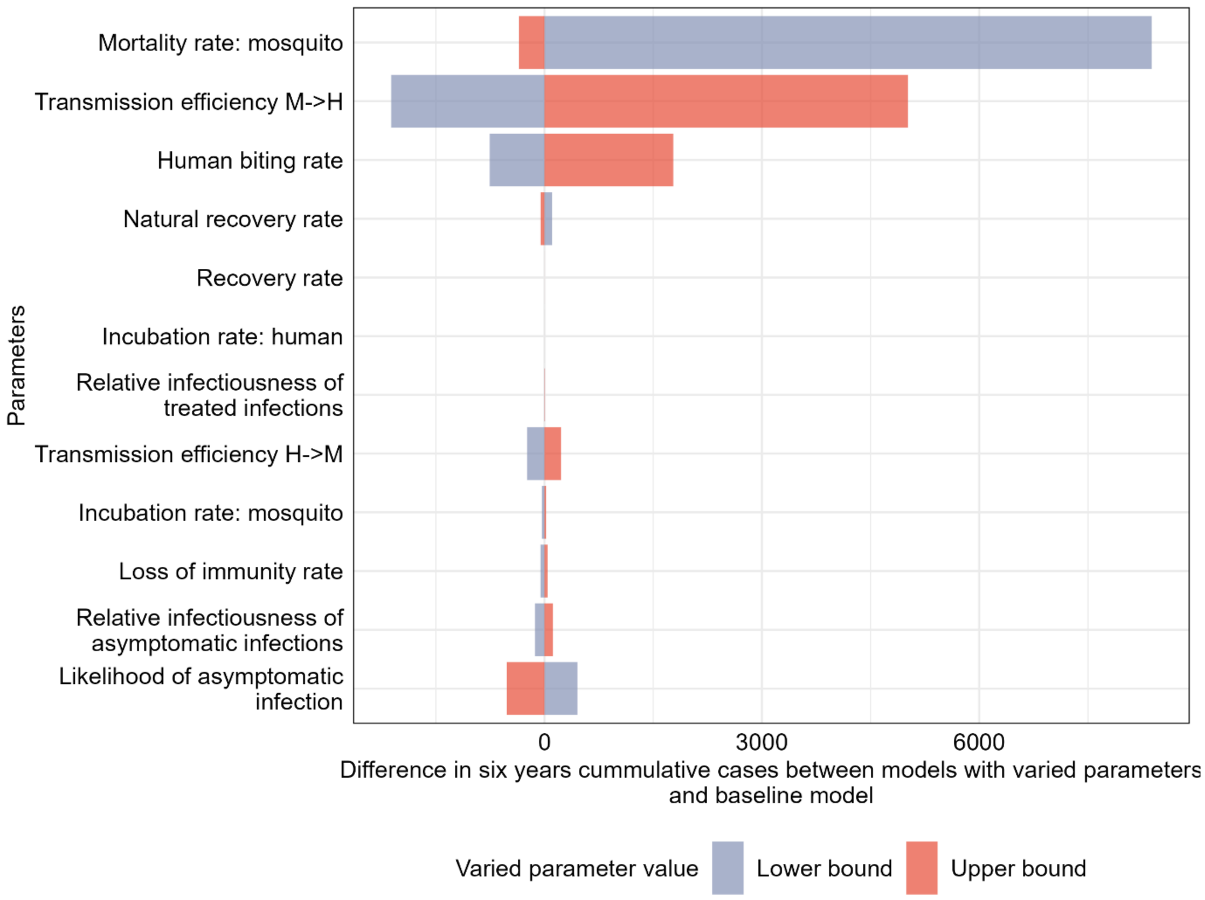


*Figure 5: The difference in cumulative cases after six years between the model with baseline (central)*

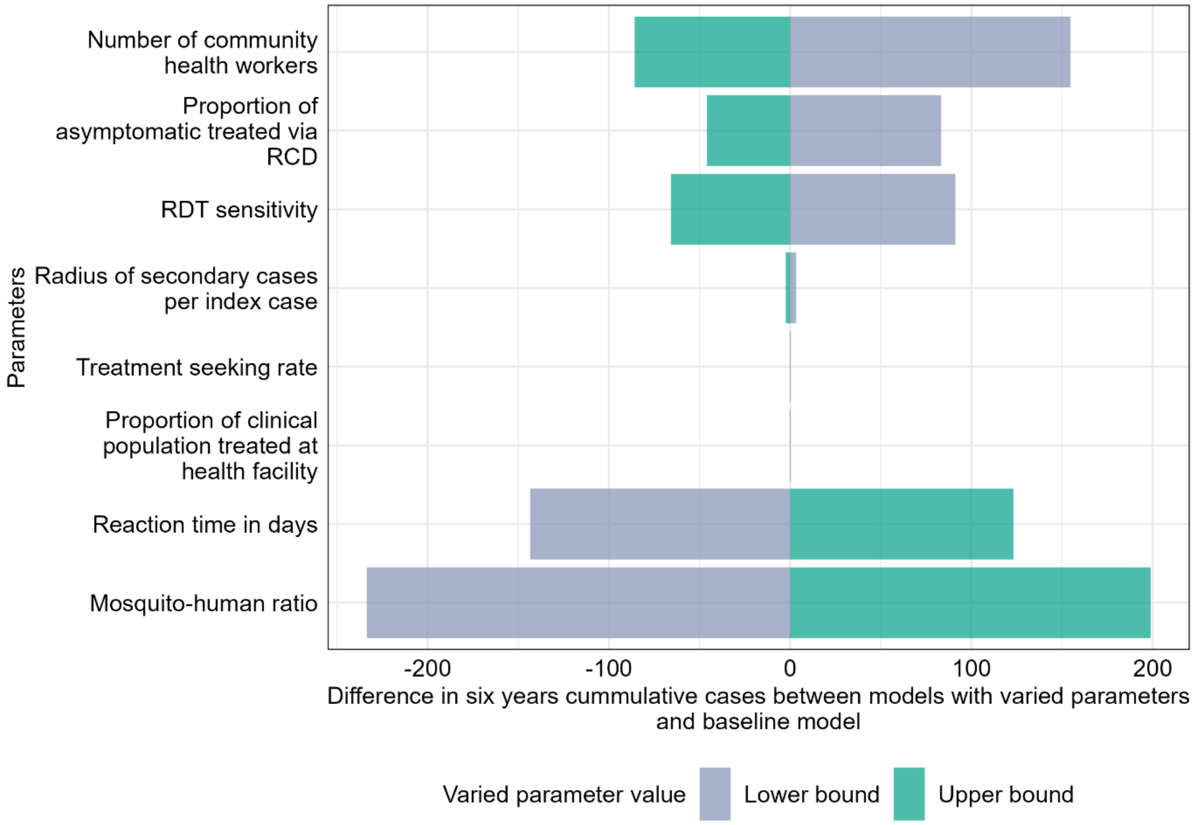


*Figure 6: Difference in cumulative cases after six years between the model with baseline (central)*

*Table 4: Sensitivity analysis results*

| term | estimate | std. error | statistic | p.value |
|---|---|---|---|---|
| (Intercept) | -11909.70 | 23773.18 | -0.50 | 0.62 |
| a | 31073.32 | 2560.11 | 12.14 | 0.00 |
| b | 81920.06 | 8543.01 | 9.59 | 0.00 |
| c | 22802.32 | 3877.44 | 5.88 | 0.00 |
| $\gamma_m$ | -7296.89 | 17448.88 | -0.42 | 0.68 |
| $\mu_m$ | -267859.22 | 23029.21 | -11.63 | 0.00 |
| $\gamma_h$ | 82653.94 | 115969.23 | 0.71 | 0.48 |
| $\tau$ | 1692.38 | 15092.49 | 0.11 | 0.91 |
| r | -1436.04 | 17707.24 | -0.08 | 0.94 |
| $\rho$ | 1735064.62 | 197274.47 | 8.80 | 0.00 |
| m | 11871.85 | 2712.60 | 4.38 | 0.00 |
| pa | -20303.76 | 7678.77 | -2.64 | 0.01 |
| $\delta$ | -280244.85 | 141993.19 | -1.97 | 0.05 |
| da | -17601.52 | 12084.19 | -1.46 | 0.15 |
| ga | -7887.72 | 15127.72 | -0.52 | 0.60 |
| $\theta$ | 32921.50 | 4389.91 | 7.50 | 0.00 |
| $\zeta$ | 16.17 | 9139.84 | 0.00 | 1.00 |
| CHW | -106.21 | 109.10 | -0.97 | 0.33 |
| radius | 7.67 | 6.38 | 1.20 | 0.23 |
| RDT sens. | -11168.53 | 7426.01 | -1.50 | 0.13 |

| | | | | |
|---|---|---|---|---|
| Reaction time | 763.78 | 589.62 | 1.30 | 0.20 |
